# regentrans: a framework and R package for using genomics to study regional pathogen transmission

**DOI:** 10.1101/2021.07.25.21261097

**Authors:** Sophie Hoffman, Zena Lapp, Joyce Wang, Evan S Snitkin

## Abstract

Increasing evidence of regional pathogen transmission networks highlights the importance of investigating the dissemination of multidrug-resistant organisms (MDROs) across a region to identify where transmission is occurring and how pathogens move across regions. We developed a framework for investigating MDRO regional transmission dynamics using whole-genome sequencing data and created regentrans, an easy-to-use, open source R package that implements these methods (https://github.com/Snitkin-Lab-Umich/regentrans). Using a dataset of over 400 carbapenem-resistant *Klebsiella pneumoniae* isolates collected from patients in 21 long-term acute care hospitals over a one-year period, we demonstrate how to use our framework to gain insights into differences in inter- and intra-facility transmission across different facilities and over time. This framework and corresponding R package will allow investigators to better understand the origins and transmission patterns of MDROs, which is the first step in understanding how to stop transmission at the regional level.

**Impact statement:** Increasing evidence suggests that pathogen transmission occurs across healthcare facilities. Genomic epidemiologic investigations into regional transmission shed light on potential drivers of regional prevalence and can inform coordinated interventions across healthcare facilities to reduce transmission. Here we present a framework for studying regional pathogen transmission using whole-genome sequencing data, and a corresponding open-source R package, regentrans, that implements these methods to streamline analyses and make them more accessible to other researchers and public health practitioners. We also discuss how these methods can be extended to study transmission in other settings.

**Data summary:** The authors confirm all supporting data, code and protocols have been provided within the article or through supplementary data files.

- The regentrans R package can be downloaded from GitHub: https://github.com/Snitkin-Lab-Umich/regentrans/
- The manuscript figures are generated from regentrans example data and can also be found on GitHub: https://github.com/Snitkin-Lab-Umich/regentrans/tree/master/vignettes/manuscript_figures
- The example data used in the package and manuscript is from BioProject accession no. PRJNA415194. The specific SRA accession numbers can be found in supplementary file S1. The metadata corresponding to these sequences can be found on the SRA Run Selector (isolate column) and as example data in the regentrans package.
- The KPNIH1 sequence was used as the reference genome (SRA accession number SRZ080789)

## Introduction

Multidrug-resistant organisms (MDROs) are a global public health threat due to limited treatment options paired with widespread global transmission [1]. Healthcare facilities in particular, where critically ill patients reside in close proximity to one another, are hotspots of MDRO transmission [2]. Furthermore, increasing evidence suggests that substantial transmission occurs not only within facilities, but also between facilities in regional healthcare networks, and that intra- and inter-facility transmission does not occur evenly across these networks [3, 4]. This observation, paired with limited resources for state and regional public health efforts, necessitates the identification of optimal intervention locations to reduce overall regional prevalence. Investigating MDRO transmission from a regional perspective can shed light on the origin and spread of MDROs, providing critical information for precision public health interventions to allocate resources to maximally reduce transmission across a region [5, 6].

Understanding where and how recent transmission is occurring is an integral first step in developing interventions to curb transmission at the regional level. A powerful tool for studying regional pathogen transmission is whole-genome sequencing, which allows us to investigate pathogen movements at very high resolution [4, 6]. Several studies have used whole-genome sequencing, sometimes paired with additional epidemiological metadata, to gain insights into locations [6, 7] and drivers [3, 4, 8] of elevated intra- or inter-facility transmission. These types of analyses have the potential to transform our public health response to MDROs if they are regularly performed at, or in collaboration with, regional public health centers. However, regional transmission analyses require careful consideration of the biological and epidemiological context, and the methods can be time consuming to implement.

Here, we aim to reduce the barrier for performing regional transmission analyses by providing a framework for studying regional pathogen transmission using whole-genome sequencing data, suggesting techniques for making analysis decisions, and presenting regentrans, an open-source R package that implements these methods. We discuss methods to study transmission within and between healthcare facilities using whole-genome sequencing data from single-colony isolates from patients, and discuss how these methods can be applied to study transmission within and between other locations such as zip codes. The methods presented here focus on studying recent transmission in a clonal set of isolates and can be applied to investigate overall transmission or transmission patterns over time, and to compare the transmission dynamics of different strains circulating in a region. We believe that these tools will help investigators more easily perform regional pathogen transmission analyses, and thus potentially guide interventions to reduce transmission.

## Investigating regional transmission patterns

Below we describe the questions, data, and methods for studying regional pathogen transmission. More details about using regentrans to implement these methods can be found in the package vignette, a step-by-step guide to using the package for an analysis.

### Questions

Our framework for studying regional pathogen transmission aims to help investigators interrogate the following questions (**Table 1**):

1. Is transmission occurring within and/or between facilities?
2. What facilities is transmission occurring within/between?
3. Have transmission dynamics changed over time?
4. Is transmission occurring along paths of higher patient/person flow?
5. Are there any observable geographic trends in prevalence/transmission?

**Table 1:** Questions regentrans can help investigate, and corresponding regentrans functions. (bold indicates main function for method)

|  | Question | Method | regentrans function | Reference |
| --- | --- | --- | --- | --- |
| Q0 | How do you choose pairwise genetic distance thresholds? | Visualize the fraction of intra-facility pairs for various pairwise genetic distances | get_pair_types(),<br>**get_frac_intra()** | [4] |
| Q1 | Is transmission occurring within and/or between locations? | Pairwise genetic distances within and between locations | **get_pair_types()** | [7] |
| Q2 | What locations is transmission occurring within/between? | Phylogenetic clustering of isolates from the same location | **get_clusters()** | [6] |
|  |  | Population-level similarity between locations | `get_genetic_flow()` | [8] |
|  |  | Number of closely related pairs within and between locations | - | - |
| Q3 | Have transmission dynamics changed over time? | Methods above but split over time | - | [7] |
| Q4 | Is transmission occurring along paths of higher patient/person flow? | Compare patient/person flow between locations to inter-location pairwise genetic distances or genetic flow (Fsp) | `get_patient_flow()`,<br>`get_pair_types()`,<br>`get_genetic_flow()`,<br>`summarize_pairs()` | [3] |
| Q5 | Are there any observable geographic trends in prevalence/transmission? | Visualize geographic distribution of prevalence and closely related pairs or genetic flow (Fsp) | `get_pair_types()`,<br>`get_genetic_flow()` | [4] |

### Data

#### Input data

Whole-genome sequences of isolates combined with corresponding metadata can be used to identify the genetic relatedness between isolates and subsequently investigate intra- and inter-facility transmission (**Figure 1**). Note that users can input data from multiple isolates from a given patient as the corresponding regentrans functions use this information in generating the output data (see the “Handling multiple isolates from a given patient” section for more details). Below we describe each component of the input data in more detail.

**Figure 1:**
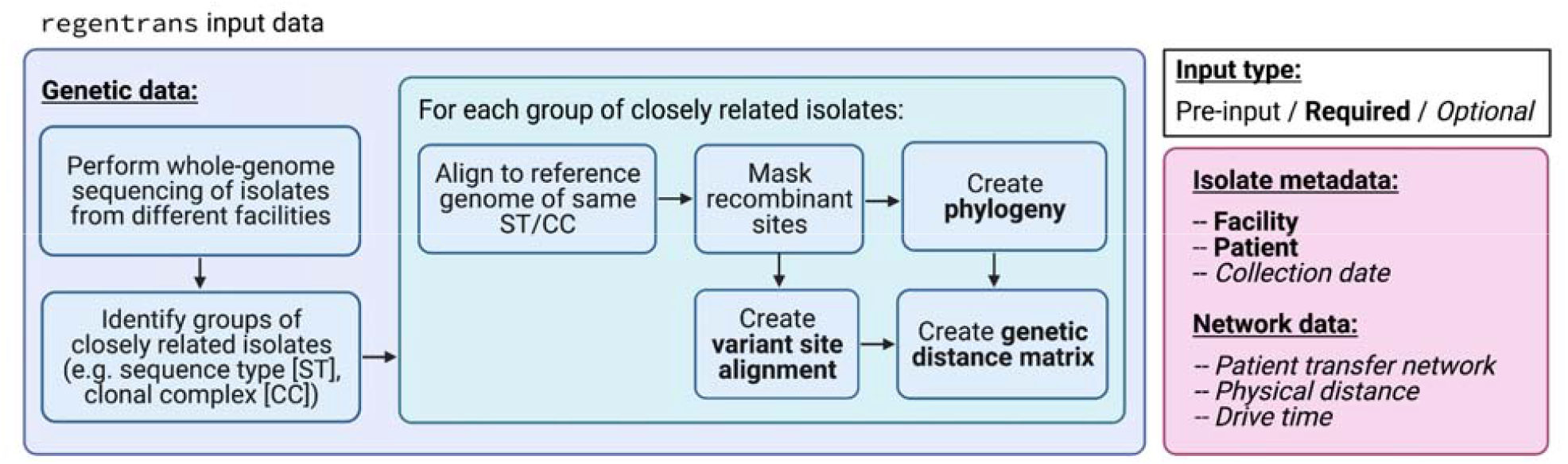
regentrans input data. Required and optional isolate genetic data and metadata for using regentrans. Figure created on BioRender.com.

##### A note on study design and sampling density

The data can come from studies such as prospective observational studies, point-prevalence surveys, and regional surveillance across different facilities in a region. While comprehensive sampling of culture or surveillance isolates from all facilities in a region is ideal, inferences can still be made using a subset of isolates and a subset of facilities. In general, users must consider how the study design, missing samples, and missing facilities may influence the results and use this to guide their interpretations. We find it useful to perform thought experiments considering what the results might look like under counterfactual scenarios where more comprehensive data was included.

##### Genetic data

The core functionality of regentrans assumes that users have whole-genome sequences of each isolate to be included in the analysis. As the methods we present here are focused on identifying recent transmission events, we suggest that they be used only on closely related isolates, e.g. ones within the same sequence type (ST) or clonal complex. However, comparisons between the transmission dynamics of different groups can be performed.

Depending on the method being used to study transmission, the genetic data required is either a phylogeny of all isolates, a genetic distance matrix, or a variant alignment. We highly recommend using a recombination-filtered variant alignment to ensure that variants included in the analysis arose from vertical gene transfer rather than horizontal gene transfer, as an assumption of the described methods is that isolates with a more recent common ancestor are more closely related at the variant level. Users can also remove variant sites known to be under positive selection. We suggest using Gubbins [9] to mask recombinant sites, IQ-TREE [10] to generate a maximum-likelihood phylogeny from the recombination-masked alignment, and the ape package [11] in R to calculate a pairwise genetic distance matrix. The genetic distance matrix can be any type of genetic distance as long as smaller numbers indicate more similarity between two isolates. Two examples of genetic distances are pairwise single nucleotide variant (SNV) distances (can be calculated with ape::dist.dna()) and patristic distances, or the sum of the branch lengths that connect two isolates on a phylogeny (can be calculated with ape::cophenetic.phylo()). While patristic distances properly account for ancestral relationships between strains, we often use pairwise SNV distances as they are easier to interpret and are the current standard with respect to transmission inference. Concordance between results using these two distances would also increase confidence in using pairwise SNV distances for interpretability.

##### Isolate metadata

For each isolate, users need to know the facility at which the isolate was collected and the patient from whom the isolate was collected. If only one isolate was collected from each patient, patient information is not required. Furthermore, isolate collection dates are needed if researchers are interested in studying transmission over time.

##### Facility-level connectedness data

Researchers can investigate the relationship between genetic distance and patient transfer between facilities, which requires a patient transfer network that minimally includes all facilities represented in the dataset. Additionally, it is possible to visualize and quantify potential geographic trends in prevalence and transmission, for which information on the physical distance, or driving time, between facilities is required.

#### Datasets used in the package and for analyses

##### Genomic data

The genomic data used for this manuscript, and included in the regentrans package, were generated from whole-genome sequences of clinical isolates obtained from 21 long-term acute care hospitals across the U.S. [4]. The data was processed as in [12]. Briefly, trimmed Illumina short reads were aligned to the KPNIH1 reference genome (BioProject accession no. PRJNA73191) using the Burrows-Wheeler short-read aligner (bwa v0.7.17) [13] and recombinant sites were masked using Gubbins v2.3.4 [9]. We used the Gubbins variant output fasta file to generate a pairwise single nucleotide variant (SNV) distance matrix using the dist.dna() function (method = ‘N’, pairwise.deletion = TRUE, as.matrix = TRUE) and a patristic distance matrix using the cophenetic.phylo() function, both in ape v5.5 [11]. IQ-TREE v1.6.12 [10] was used to generate a whole-genome phylogeny of all isolates. All non-recombinant variants, including variants that may have been under positive selection, were included when calculating pairwise SNV distances and generating the phylogeny. For all analyses, the data was subset to include only ST258 isolates. Sequence types were determined using Kleborate v0.4.0 [14].

##### Isolate metadata

We used information on the facility and the patient the isolate was taken from, as well as the isolate collection date.

##### Patient transfer data

Aggregate patient transfer data from all hospitals in the state of California was used to calculate paths of maximum patient flow between the 11 Los Angeles area facilities in our dataset. The data and methods are described in [4].

##### Geographic data

The example geographic data was de-identified by adding random horizontal, vertical, and rotational shifts using the R package tangles v0.8.1 [15].

## Methods

### Q0: How do you choose pairwise genetic distance thresholds?

Several of the methods discussed below rely on interpreting, comparing, or thresholding pairwise genetic distances between isolates to make inferences. It is generally understood that small pairwise distances between isolates implies recent transmission [16–18], but that this method is not entirely accurate due to within-host evolution and variable mutation rates [19, 20]. To identify recent transmission pairs using pairwise distances, investigators must choose a threshold to determine what pairs are considered closely related [16–18]. The threshold for “closely related” depends on the pathogen mutation rate and genome size, and the setting. Within a given time period, a higher mutation rate or a larger genome size will lead to higher pairwise distances. Additionally, the mutation rate of pathogens in outbreak settings is often higher than endemic settings [21]. Thus, for a given pathogen, closely related isolate pairs in an outbreak setting will likely have a higher pairwise genetic distance than closely related pairs in an endemic setting. For this reason, knowledge of the epidemiologic context of the isolates, and the species or sequence type itself, is essential for correctly interpreting pairwise genetic distance distributions. Here, we discuss two methods for identifying reasonable pairwise SNV distance thresholds. Similar methods can be used to identify thresholds for other pairwise genetic distance metrics, such as pairwise patristic distances.

One way to choose a pairwise SNV distance threshold is to use the genome length and mutation rate. For instance, in the context of the dataset we use here, *K. pneumoniae* ST258 isolates from an endemic setting, we could calculate a pairwise SNV distance threshold of 11 SNVs based on the KPNIH1 reference genome length of 5,394,056 base pairs and a mutation rate of 1.03e-6 mutations per base pair per year [22] (2 isolates * 5,394,056 bases * 1.03e-6 mutations per base per year per isolate). Mutation rates can be estimated for measurably evolving populations with a temporal signal [21–23]. However, mutation rate estimates are less reliable for lineages with high among-lineage rate variation or phylogenetic and temporal clustering [24], and are unreliable for datasets with no temporal signal [24]. Therefore, it is often difficult or impossible to reliably calculate the evolutionary rate of the pathogen for the particular dataset being used, and more general estimates of mutation rate may not be translatable.

If a mutation rate cannot be reliably calculated, or to perform a sensitivity analysis, one way to identify potential pairwise SNV distance thresholds is to use the assumption that pairs of individuals from the same facility will be enriched for recent transmission relative to pairs from different facilities. In particular, this method requires you to plot the fraction of isolate pairs from the same facility for increasing pairwise SNV distances. A decrease in the fraction of intra-facility isolate pairs at a given SNV distance suggests a potentially reasonable threshold. Performing this analysis on our data indicated that using SNV distance thresholds of ≤ 10 and ≤ 6 appear to be reasonable (**Figure 2**). We chose to use these two thresholds for our sensitivity analysis as they are more directly supported by our data compared to the more general mutation rate analysis using a mutation rate that was not calculated from the same dataset. However, in this case the results would likely be similar as the mutation rate analysis suggests using a threshold of 11 SNVs, which is similar to the 10 SNV threshold we use here.

**Figure 2:**
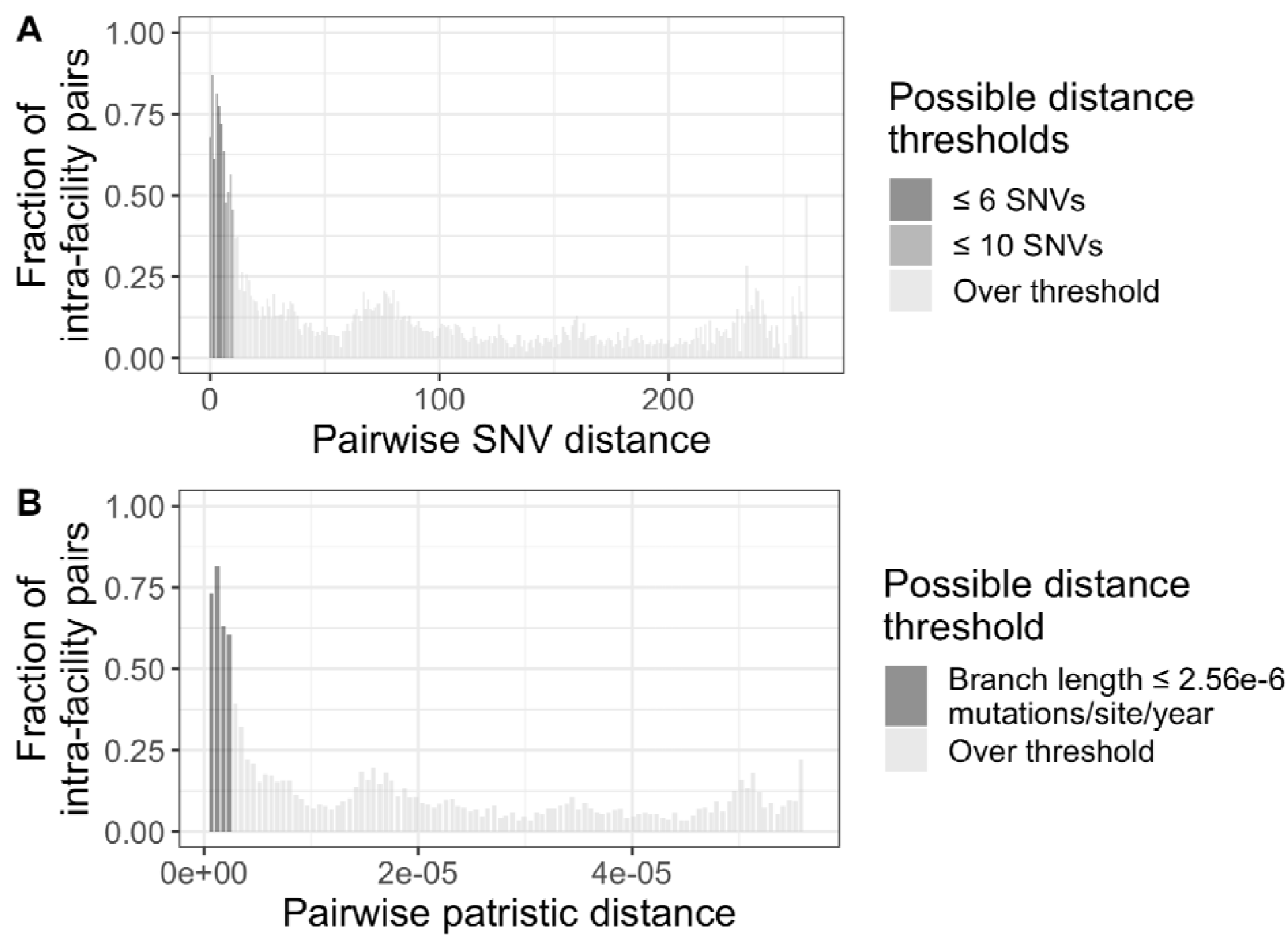
Data-specific method for choosing pairwise genetic distance thresholds. **(A)** Fraction of intra-facility pairs for various pairwise single nucleotide variant (SNV) distances. **(B)** Fraction of intra-facility pairs for various pairwise patristic distances grouped into 100 bins. These plots can help identify drops in intra-facility pair fractions that may indicate a reasonable pairwise distance threshold, assuming intra-facility transmission is more common than inter-facility transmission. Note that it may be difficult to clearly identify a large drop at any given threshold; therefore, we recommend performing sensitivity analyses with several thresholds. Furthermore, users should consider the tradeoff between sensitivity and specificity when deciding what thresholds to choose. This data can be generated using the get_frac_intra() function.

When performing analyses where choosing a pairwise SNV distance threshold is necessary, we suggest that investigators evaluate the robustness of their results by comparing their findings for different pairwise SNV distance thresholds. Here, we compare the results using a threshold of 6 and 10, but a wider range of values can be used, particularly in instances with more uncertainty about what threshold should be used.

### Q1: Is transmission occurring within and/or between facilities?

One of the first questions an investigator might ask about isolates collected from a certain region is if transmission is occurring within particular facilities and/or between facilities. Inspecting pairwise genetic distances between all isolates can provide information about the extent of recent transmission both within and between facilities, which will often manifest as an enrichment in closely related isolate pairs between patients (i.e. isolate pairs with small pairwise genetic distances; see note above on what to consider closely related). In our example dataset, we observe an enrichment in both closely related intra-facility pairs and inter-facility pairs (pairwise SNV distance of ≤ 10 or ≤ 6), suggesting that recent transmission is occurring both within and between facilities (**Figure 3**).

**Figure 3:**
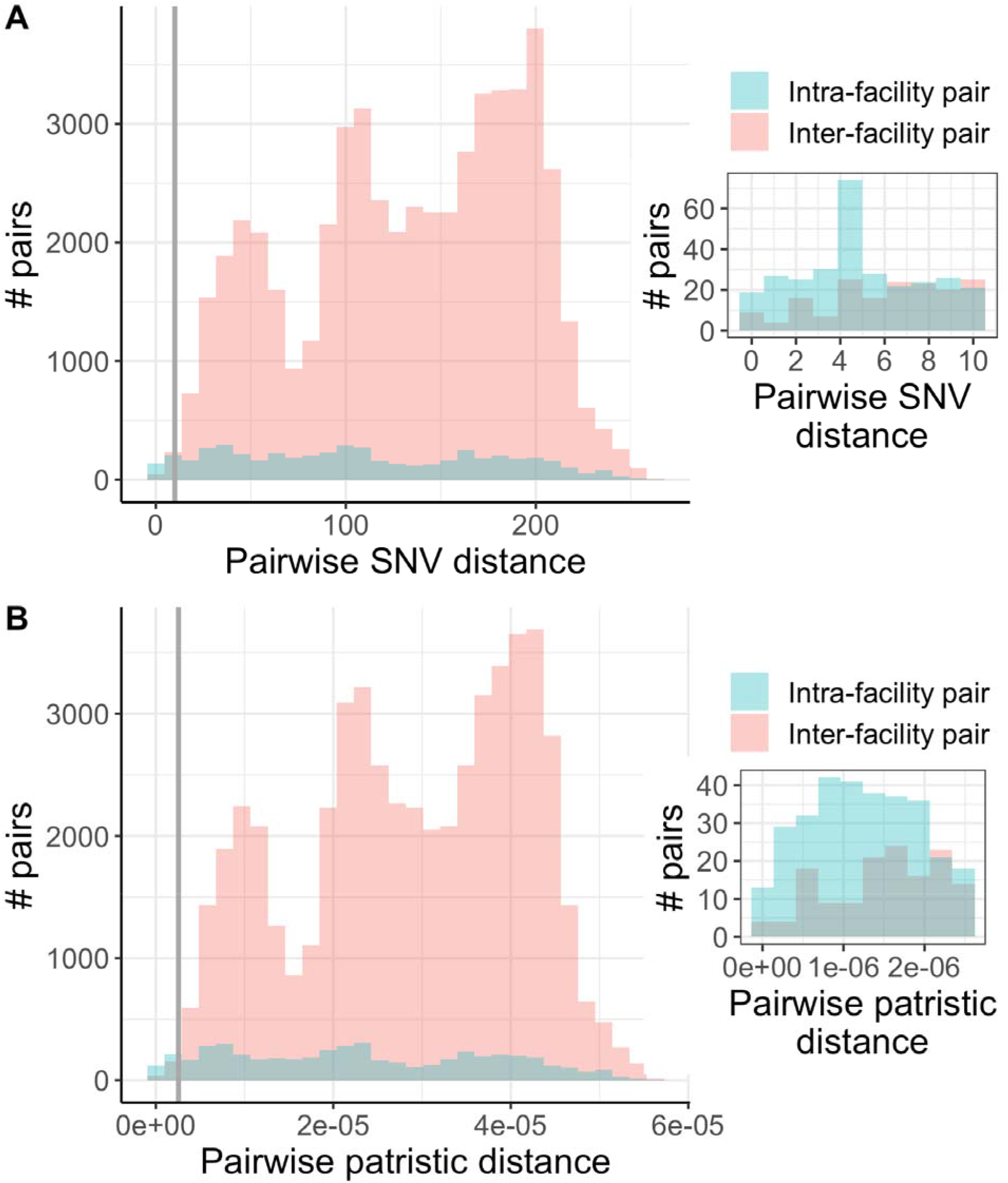
Pairwise genetic distances between facilities suggest recent intra- and inter-facility transmission. **(A)** Pairwise single nucleotide variant distances. Grey line is at a pairwise SNV distance of 10, and the inset shows all pairs with a pairwise SNV distance less than or equal to this threshold, which we consider indicative of recent transmission. See Q0 on genetic distance thresholds in the main text for more details on how we chose these thresholds.. **(B)** Pairwise patristic distances. Grey line is at a pairwise patristic distance of 2.56e-6 mutations per site per year, and the inset shows all pairs with a pairwise patristic distance less than or equal to this threshold. These plots indicate that transmission is likely occurring both within and between facilities, due to small pairwise genetic distances for both intra- and inter-facility pairs. Data generated using the get_pair_types() function.

One limitation of the pairwise genetic distance method for investigating transmission is that it may miss transmission events occurring over longer time periods when there is, for instance, sparse sampling and more evolutionary divergence between isolates. Another limitation is that researchers must define a threshold for what is considered closely related, and different thresholds may lead to different conclusions. Therefore, we recommend performing a sensitivity analysis across several pairwise genetic distance thresholds and genetic distance metrics to investigate the robustness of the results to different thresholds.

### Q2: What facilities is transmission occurring within/between?

Once we have investigated the extent of transmission occurring within and between facilities on a general scale, we can dig deeper into identifying certain facilities and facility pairs with closely related isolates. Intra-facility transmission can be investigated using phylogenetic context, and inter-facility transmission can be investigated by taking a population-level approach computing genetic flow between facilities. These methods are both threshold-free. Intra- and inter-facility transmission can also be investigated by identifying probable recent transmission events between facilities, which requires a pairwise genetic distance threshold to be defined.

#### Investigating intra-facility transmission using the phylogeny

One way to investigate the extent of intra-facility transmission is to identify maximum subclades that all originate from distinct patients from the same facility and quantify the size of these clusters. Larger clusters indicate more intra-facility transmission, as those isolates are all more closely related to one another than to isolates from other facilities. Furthermore, the range of sampling dates of each cluster can provide insight into whether the cluster is from an intra-facility outbreak (smaller date range) or from sustained intra-facility transmission (larger date range). In our dataset, we see that some facilities (e.g. facility D) exhibit extensive intra-facility transmission as evidenced by large cluster sizes, while others exhibit relatively little intra-facility transmission (e.g. facility K) (**Figure 4**). All large clusters also have large date ranges, suggesting that they are from sustained intra-facility transmission rather than a shorter-term outbreak. However, it is also important to note that, under these circumstances of sustained transmission, isolates within a cluster may be distantly related, particularly if there are few exportation events from that facility to other facilities (e.g. if the facility is more geographically isolated). This may be considered a strength of the method if users are interested in investigating transmission across longer timescales.

**Figure 4:**
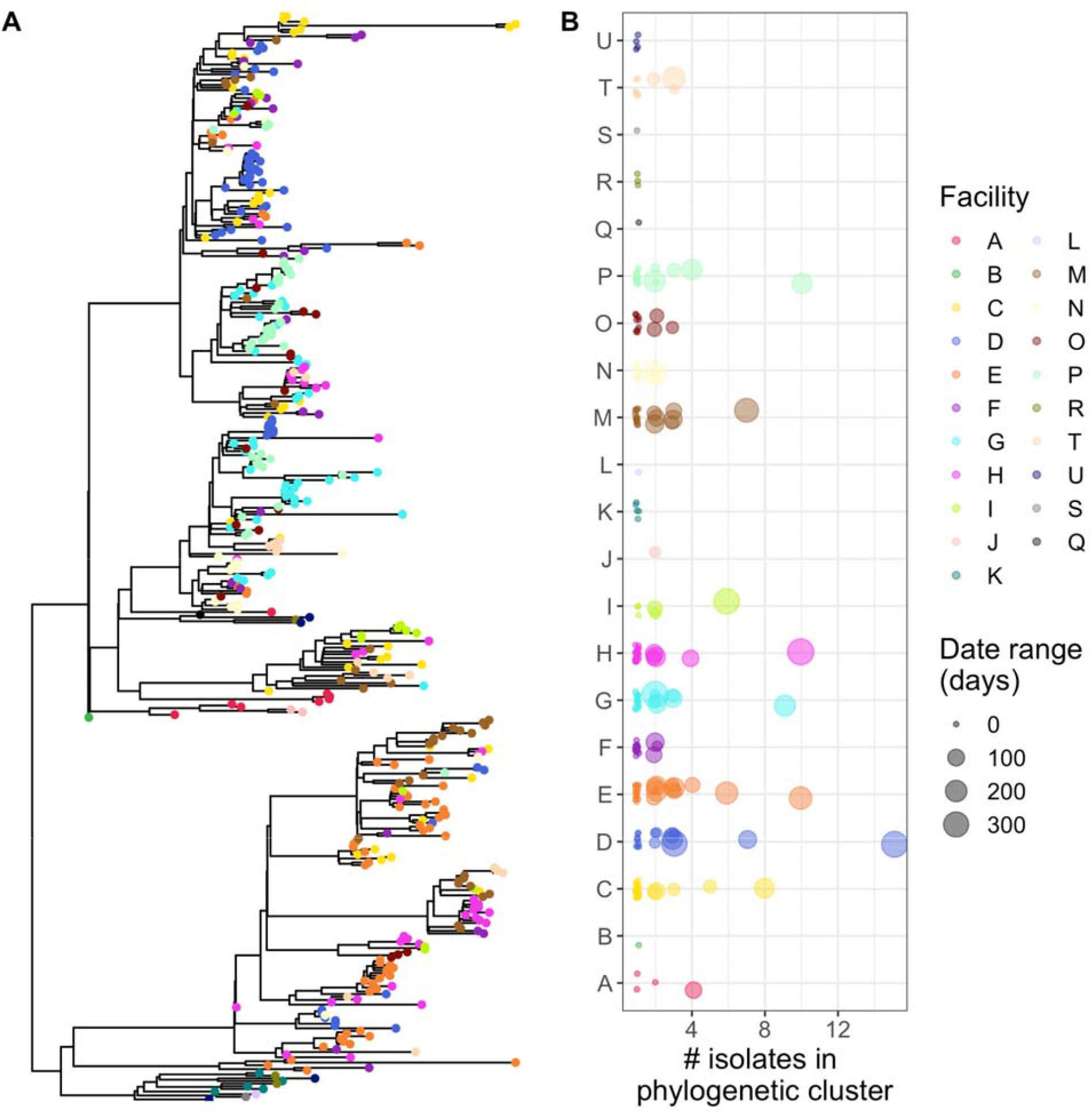
Clusters of isolates from the same facility suggest intra-facility transmission. **(A)** Mapping isolate location on the phylogeny provides a visual for the extent of clustering by facility. Here we can see clustering of isolates from the same facility in several subclades of the phylogeny. **(B)** Quantification of the size of phylogenetic clusters from a single facility using get_clusters(). The size of the points in panel B can provide insight into whether large clusters are from an intra-facility outbreak (smaller point size) or sustained intra-facility transmission (larger point size). Points in panel B are jittered for visualization purposes.

There are a few key limitations to investigating intra-facility transmission based on phylogenetic cluster size that must be considered when interpreting results. First, cluster sizes may be biased when there is skewed sampling across facilities [8], where larger clusters may be identified for over-represented facilities and smaller clusters for under-represented facilities. For instance, if unsampled strains from under-represented facilities were to be included, they may increase the cluster size of pre-existing clusters from that facility or decrease the size of clusters from another facility by being interspersed among those samples. Therefore, we recommend careful consideration of potential sampling bias prior to comparing cluster sizes across facilities. Second, if there is frequent intra- and inter-facility transmission, recent transmission events between facilities may not be identified as these clusters will be broken up by recent inter-facility transmission events. We provide the option to reduce the cluster “pureness” to attempt to accommodate this limitation. Finally, the clustering method does not directly allow investigators to assess the extent of inter-facility transmission. The genetic flow and pairwise genetic distance methods described below address these limitations of the cluster size method.

#### Genetic flow between facilities

Identifying the extent to which variants are shared among isolates at different facilities by calculating genetic flow between facilities (Fsp) [8] provides a threshold-free population-level approach to investigating the extent of inter-facility transmission. Fsp is a measure of population-level similarity between facilities, where an Fsp value closer to 0 indicates more genetically similar populations and a value closer to 1 indicates more genetically different populations. This method is particularly useful in endemic scenarios when the relationship between individual isolates may be relatively diffuse due to frequent patient transfer over time. Using our dataset, we found that certain facility pairs (e.g. facilities G and P) have more genetic flow between them compared to others, indicating that there is likely more transmission between those facilities (**Figure 5**). Facilities that have high Fsp compared to other facilities (e.g. J) have populations that are genetically distant from all other facilities, which indicates that they are likely less involved in the inter-facility transmission network. Note that one limitation of using Fsp is that small sample sizes may overestimate genetic differences, although this is less of a concern when using many variant sites [27].

**Figure 5:**
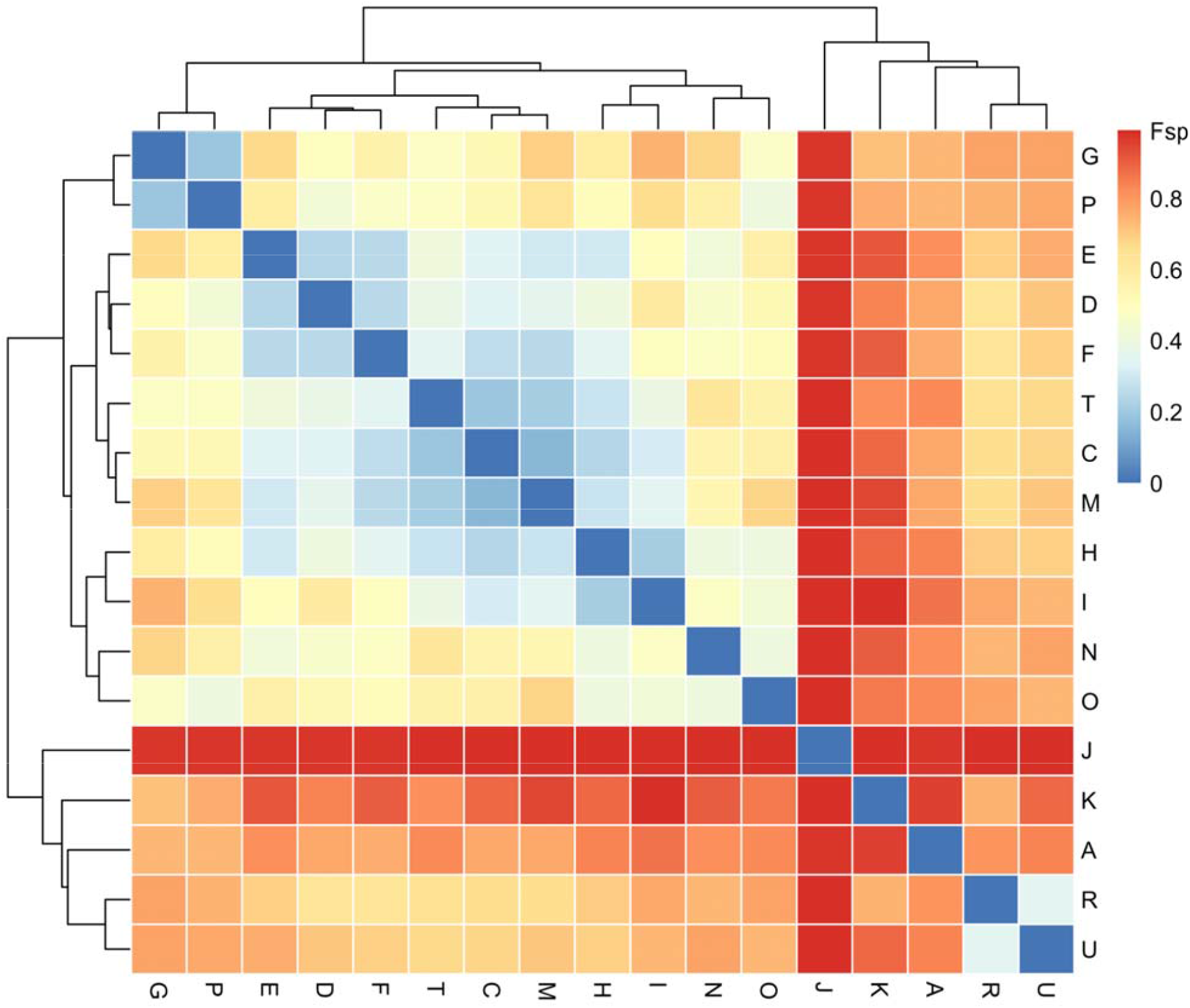
Some facility pairs have similar populations, indicating potential transmission between them. Genetic flow. (Fsp) was calculated using the get_genetic_flow() function in regentrans. Rows and columns are facilities. Lower Fsp indicates more similar populations and thus more putative transmission. At least two isolates are required from a facility to perform the Fsp calculation as within-facility variation must be calculated, so facilities with only one isolate have been removed. The heatmap is clustered based on Fsp values.

#### Pairwise genetic distance threshold

Using a pairwise genetic distance threshold, the number of closely related pairs within and between facilities can be determined and used to identify facilities and with more or less putative spread. For instance, we observed that not only are there differing extents of spread at different facilities, but some facilities have more closely related intra-facility pairs than inter-facility pairs (e.g. facility D), while other have more even amounts of closely related intra- and inter-facility pairs (e.g. facility H) (**Figure 6A**). These findings are generally concordant with our findings from the phylogenetic clustering and genetic flow analyses as well (e.g. more intra-facility transmission in facility D, isolates from facility H being related to isolates from other facilities). Furthermore, as there are limitations to choosing genetic distance cutoffs, we highly recommend doing a sensitivity analysis by choosing several different thresholds and using different genetic distance metrics to see how robust the results are to these changes.

**Figure 6:**
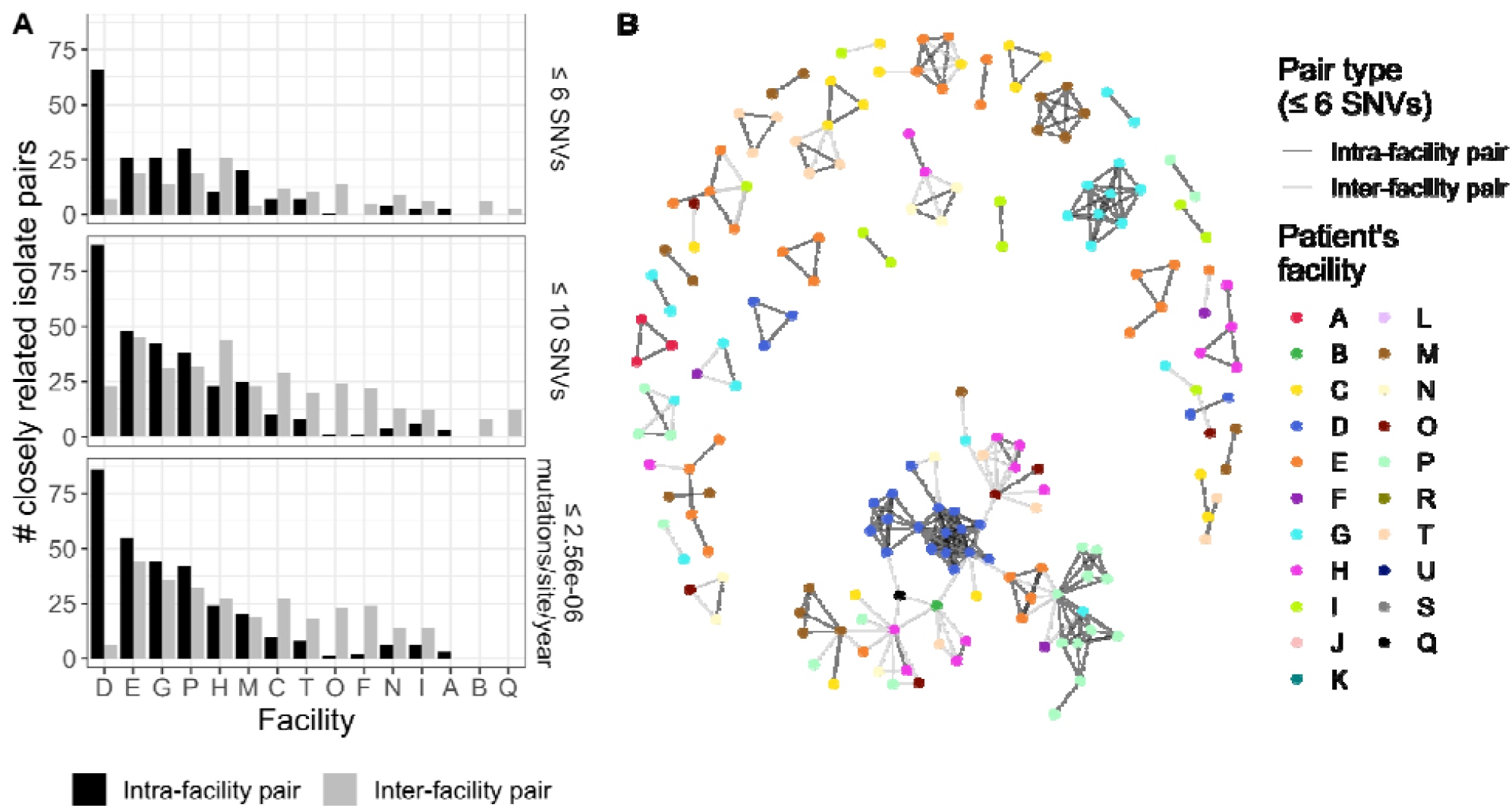
Some facilities have many closely related isolates, indicating potential intra- and inter-facility transmission. **(A)** Number of closely related isolate pairs for different genetic distance metrics and thresholds. Only facilities with at least one pair of closely related isolates are shown. **(B)** Network of patient connectedness. Patient nodes are connected by edges if they share an isolate with a pairwise SNV distance ≤ 6.

One limitation of visualizing the number of closely related pairs within and between facilities is that it does not provide insight into the potential transmission networks among patients in these pairs. For instance, we do not know how often each patient isolate is included in these pairs, or whether the pairs are closely related to each other. This question can be probed by visualizing the connectedness of patients with closely related isolates. For instance, in our data we see that there is an isolated network of closely related isolates from facility G, as well as a network of closely related isolates from facility D that is connected to isolates from other facilities (**Figure 6B**).

The phylogenetic clustering, genetic flow, and pairwise genetic distance methods provide complementary information related to intra- and inter-facility transmission. We suggest evaluating the strengths and limitations of each method with respect to the dataset being used and the questions of interest to determine which method(s) to use for a given application. Concordance between methods provides additional confidence in the results.

### Q3: Have transmission dynamics changed over time?

All of the methods described above can be applied to discrete time windows to gain insight into whether transmission dynamics have remained stable or changed over time. For instance, in an outbreak setting we observed an increase in intra-facility transmission followed by an increase in inter-facility transmission [7]. In an endemic setting, these trends may remain more stable over time. In our data, we observe an increase in the total number of pairs from 2014 to 2015, but no change in the distribution of closely related intra- or inter-facility isolate pairs (**Figure 7**).

**Figure 7:**
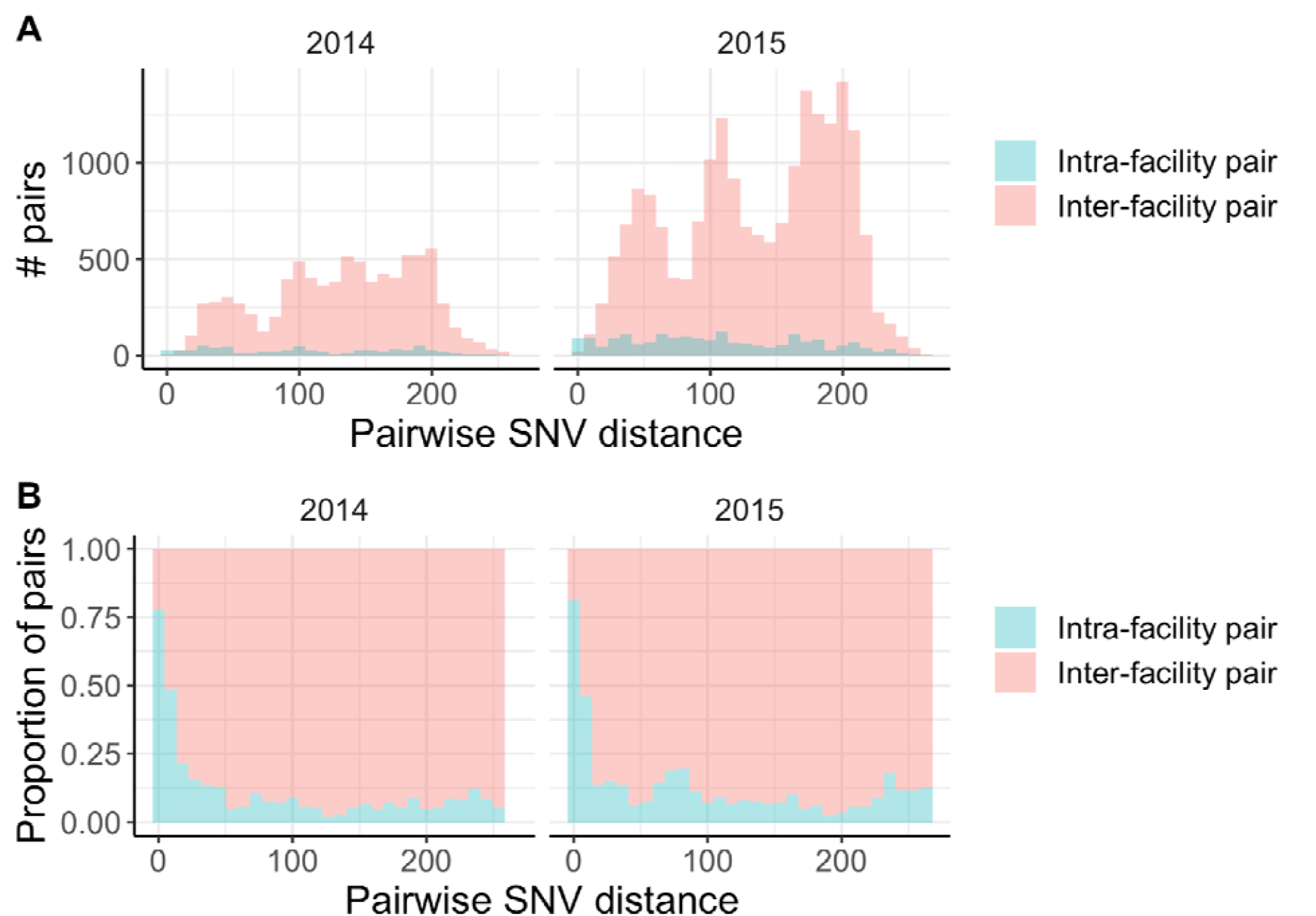
The pairwise SNV distance distribution does not change over time. (A) Count of pairwise single nucleotide variant (SNV) distances faceted by year. (B) Fraction of intra- vs. inter-facility pairwise SNV distances faceted by year. Trends are similar across both years.

### Q4: Does transmission correlate with patient/person flow?

In addition to only using genomic information to study transmission, inter-facility transmission can be studied in the context of patient flow between facilities. While sometimes investigators may have access to patient-level information regarding prior facility exposures, this information is often not available. In this case, aggregate patient transfer data can be used to study the relationship between patient flow and transmission. The simplest way to do this is to determine whether there is a relationship between direct flow between facilities and either the number of closely related pairs defined by pairwise SNV distances, the actual values of pairwise SNV distances, or genetic flow (Fsp). To take into account potential indirect transfers that may influence transmission, a more complex algorithm can be used to identify paths of maximum patient flow between facilities, and then this can be compared to metrics of genomic relatedness [4]. These analyses can provide insight into whether patient flow may be driving transmission between facilities. For instance, when subsetting our data to the 11 Los Angeles area facilities for which we have information on inter-facility patient transfer events, we observed a negative correlation between patient flow and genetic flow (Fsp), indicating that facilities connected by more patient flow often have more similar populations (**Figure 7A**). We also observed a negative correlation between patient flow and minimum pairwise SNV distance between facilities (**Figure 7B**), and a positive correlation between patient flow and the number of closely related isolate pairs between facilities (**Figure 7C**). Taken together, this suggests that patient flow may, in part, drive inter-facility transmission. However, it is important to note that these correlations are driven by facilities with high and low patient flow. This indicates that genomic data adds information about regional genomic transmission that is not captured by the static patient transfer network, particularly for facility pairs with intermediate patient flow between them.

### Q5: Are there any observable geographic trends in prevalence/transmission?

Finally, it is often useful to visualize the geographic distribution of closely related isolates. This can provide insight into whether inter-facility transmission is concentrated in a certain geographic region, or is more diffuse. For instance, we can see in our data that facilities that are geographically more proximate tend to have more transmission between them (**Figure 9**), as indicated by a positive correlation between geographic distance and genetic flow (Fsp; **Figure 9B**) and a negative correlation between geographic distance and number of closely related isolate pairs for a given facility pair (**Figure 9D**). Interestingly, we do not see a correlation between geographic distance minimum pairwise SNV distance between facilities (**Figure 9C**), possibly because this metric is more susceptible to noise.

**Figure 8:**
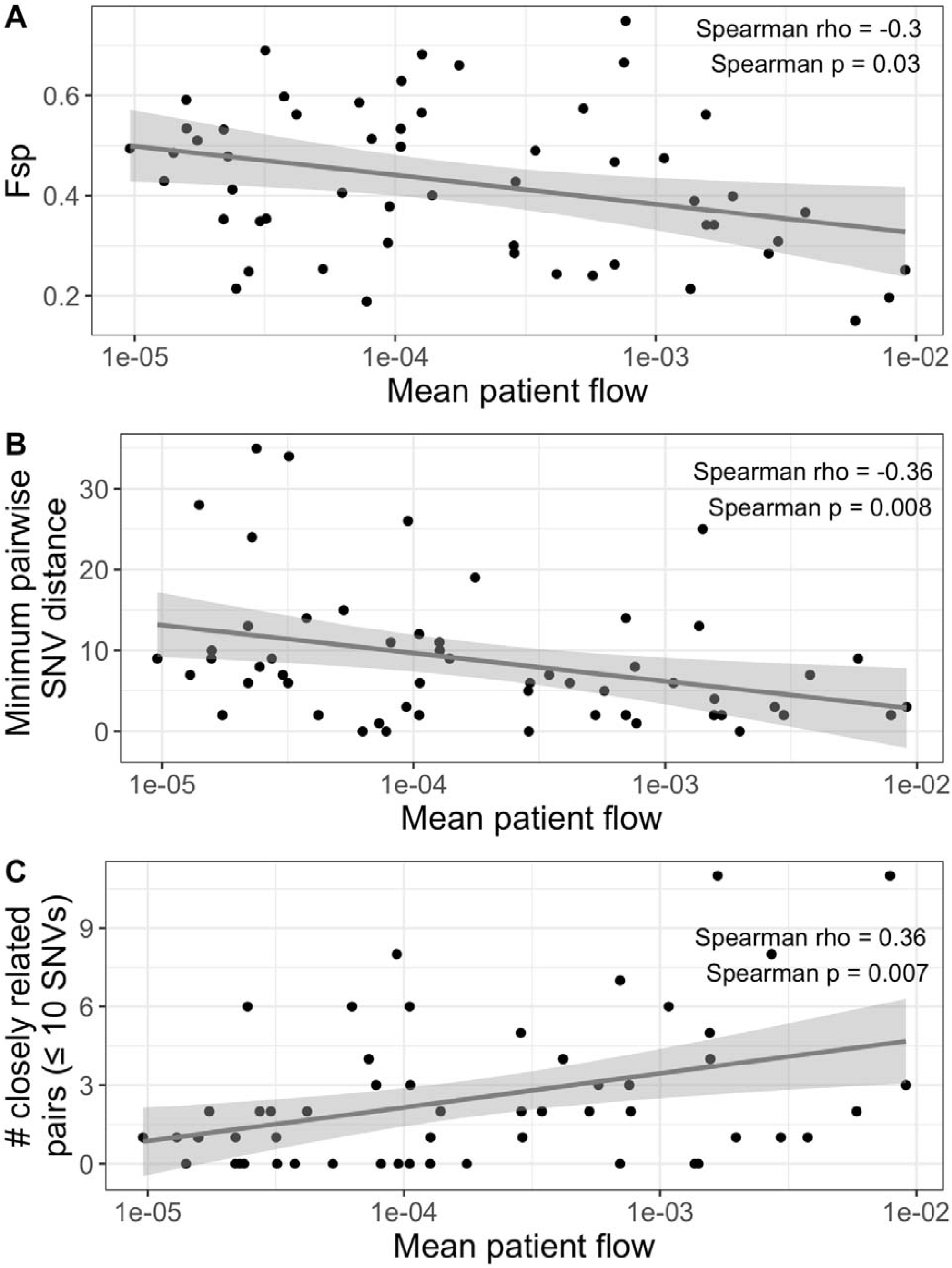
Facilities with more patient flow tend to have more similar *K. pneumoniae* populations. **(A)** Patient flow and genetic flow (Fsp) are negatively correlated. (**(B)** Patient flow and the minimum pairwise single nucleotide variant (SNV) distance (i.e. the SNV distance between the most closely related isolates) are negatively correlated. **(C)** Patient flow and number of closely related isolate pairs are positively correlated. Patient flow is the path of maximum patient flow. For this analysis we considered indirect transfers as long-term acute care hospitals are often not connected by direct transfers, but rather are connected by transfers to an intermediate facility such as an acute care hospital. Mean patient flow was calculated as the mean of the two directional patient flow metrics between two facilities. Lines were plotted using ggplot::geom_smooth() with the ‘lm’ method.

**Figure 9:**
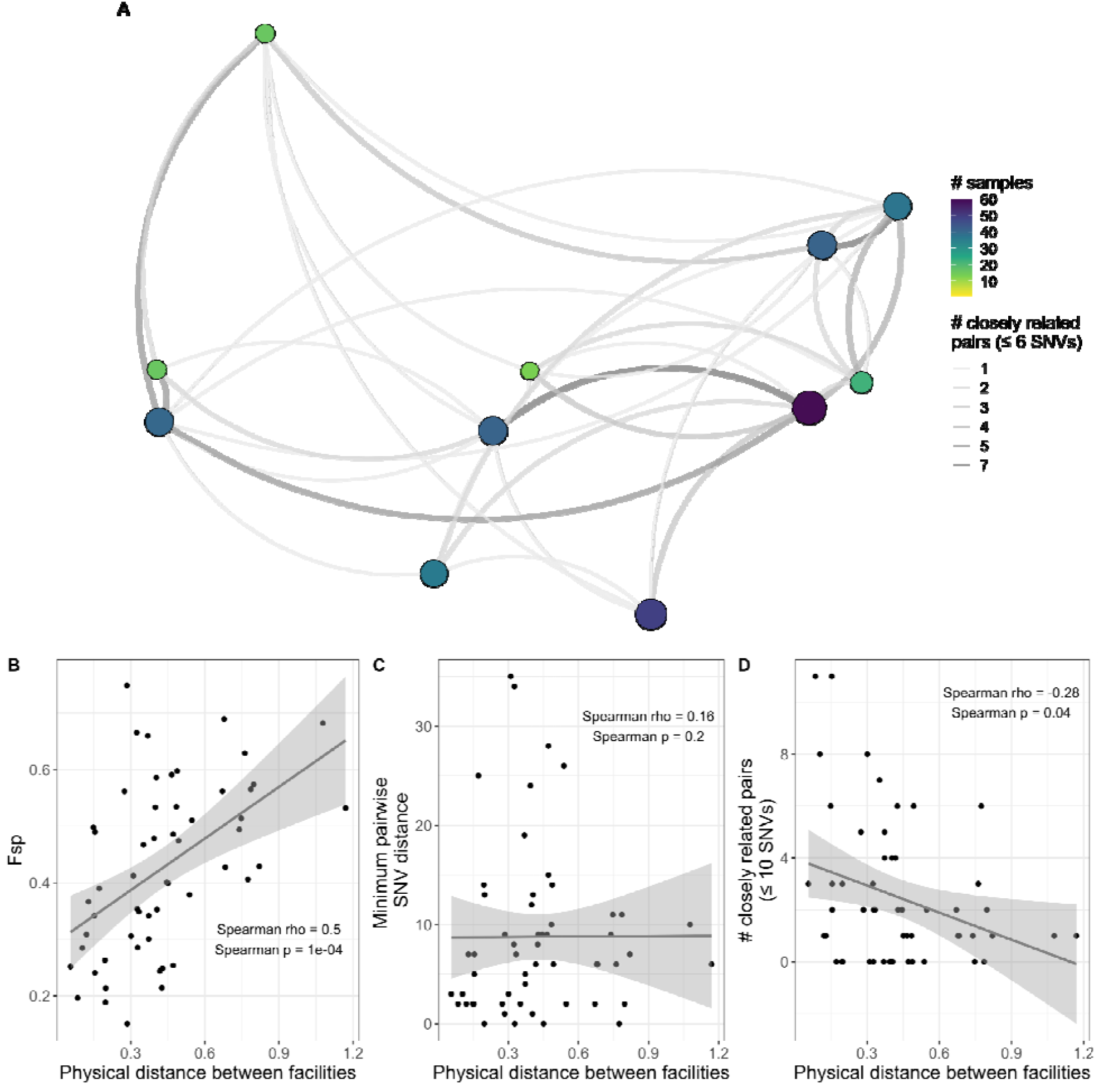
Geographically close facilities are often connected by closely related isolate pairs. **(A)** Facilities are located as they are geographically in space but latitude and longitude are de-identified by horizontal, vertical, and rotational shifts. The smaller single nucleotide variant (SNV) threshold was chosen for visualization purposes. Size and color of points indicates sample size, size and color of edges indicates number of closely related pairs. **(B)** Physical distance between facilities is positively correlated with genetic flow (Fsp). **(C)** Physical distance between facilities is not correlated with minimum pairwise SNV distance. **(D)** Physical distance between facilities is negatively correlated with number of closely related isolate pairs (≤10 SNVs). The larger SNV threshold was chosen to have a wider distribution of number of closely related isolate pairs. Physical distance was calculated as the shortest distance between the points of latitude and longitude for the facility pair. Lines in panels B, C, and D were plotted using ggplot::geom_smooth() with the ‘lm’ method.

## Package details

### Usage

The `regentrans` package provides three main functions for investigating genetic similarity between intra- and inter-facility isolates in a dataset:

1. Identifying clusters of isolates originating from the same facility (`get_clusters()`),
2. Identifying intra- and inter-facility pairwise similarity between isolates (`get_pair_types()`).
3. Calculating population-level similarity between facilities (`get_genetic_flow()`).

The corresponding output of these functions, sometimes in conjunction with other functions, can then be used to answer the questions described above. The required and optional inputs to each function can be found in **Table 1**. Many functions require a phylogenetic tree read in by ape::read.tree() and/or a genetic distance matrix that can be calculated using ape::dist.dna(), which requires a DNAbin object input that can be read in using ape::read.fasta(), or ape::cophenetic.phylo(), which requires a phylogenetic tree input.

Our introductory vignette provides examples of how to read in data, use each function, and plot the corresponding output for interpretation.

### Underlying methods

Please refer to the references noted in **Table 1** for a detailed description of the methods and algorithms used in the underlying functions [3, 4, 6, 7, 25].

### Handling multiple isolates from a given patient

Inclusion of multiple samples from an individual is desirable to capture accumulated genetic diversity; however, this requires careful consideration as it is important to ensure that observed similarities are not due to similarities within a given patient, but rather between different patients. For each of the three methods for investigating intra- and inter-facility genetic similarity, we first define unique patient-facility combinations to ensure that we capture events where a patient has transferred from one facility to another. However, we handle multiple isolates from the same patient and the same facility differently for each method. We describe these differences below. Also note that if users would like to process the data in a different or more nuanced way, they may do so prior to inputting the data into the function.

#### Phylogenetic clustering

Prior to identifying phylogenetic clusters of isolates, we subset to include only one isolate per patient per facility to ensure that all intra-facility clusters contain only one isolate per patient.

#### Intra- and inter-facility pairwise genetic distances

For each pair of unique patient-facility combinations, we include only the most closely related isolate pair.

#### Between-facility genetic similarity

As Fsp only accommodates biallelic sites, we first subset to include only biallelic sites. Then, for each patient-facility combination, we merge all sequences into a “meta-sequence” where, for each variant position the population-level minor allele is prioritized over the population-level major allele. In other words, all of the minor alleles for a patient-facility combination are merged into one sequence. This meta-sequence is used to calculate Fsp values between facilities. This method allows us to capture all minor alleles for a given patient within a given facility without biasing the population-level prevalence of a minor allele by including multiple isolates from the same patient.

### Package implementation

regentrans is implemented in R [28] and is available on GitHub (https://github.com/Snitkin-Lab-Umich/regentrans). Our package depends on several other packages including tidyverse [29] packages (dplyr and tidyr), ape [11], phytools [30], igraph [31], and future.apply [32]. The ggplot2 [29], ggtree [33], ggraph [34], and pheatmap [35] packages are used in the vignette for plotting.

## Additional possible uses

While our expertise in studying regional transmission largely lies in investigating bacterial pathogen transmission within and between healthcare facilities, the methods implemented in regentrans can be used for many additional applications. Firstly, they are generalizable to the analysis of not only other bacterial pathogens (e.g. MRSA), but also other organisms including fungal (e.g. *Candida auris*) and viral (e.g. SARS-COV-2) pathogens, and even non-pathogenic microbes. Additionally, rather than investigating transmission between facilities, users could investigate transmission between different zip codes, different rooms or wards within a hospital, or even transmission between patient and environmental sources. As one example, Popovich and Thiede *et al*. [36] identified transmission signatures within a large urban jail by comparing pairwise SNV distances of community-onset MRSA to MRSA acquired within the jail.

## Cautionary notes on interpretation

It is important to emphasize that there are several limitations to the methods we describe here. First, none of these methods include the use of epidemiological data to confirm or corroborate putative transmission links. Therefore, while we can gain useful insight into the likely extent of transmission within and between facilities, we cannot understand the nuances of actual transmission events. If epidemiological data is available, we highly recommend incorporating this information into the analysis to provide further insights into putative transmission pathways (examples: [6, 36]). Additionally, as mentioned above, for methods where choosing a threshold of genomic relatedness is required, care in choosing the threshold and investigating the sensitivity of the threshold on your interpretation of the results is warranted as the results may change drastically depending on what threshold is chosen [37]. Finally, the sampling schemes or time frames used in the study can influence the output of the methods presented here [38, 39]. Therefore, as always, the strengths and limitations of the dataset being used must be considered carefully when interpreting the results.

## Conclusion

Investigating regional pathogen transmission can provide insight into transmission dynamics and guide infection prevention and control. Here we provide a framework for studying regional pathogen transmission within and between healthcare facilities using whole-genome sequencing data, and implement these methods in the easy-to-use R package regentrans. regentrans allows users to interrogate the transmission dynamics of pathogens using various metrics of genomic relatedness, including genetic-threshold and threshold-free approaches. Using several complementary methods to investigate intra- and inter-facility transmission allows investigators to gain a better understanding of the robustness of their findings and provide different insights into transmission dynamics in the region of interest. Therefore, we believe that this tool will be a useful resource for researchers and public health practitioners interested in investigating regional pathogen transmission.

## Data Availability

The authors confirm all supporting data, code and protocols have been provided within the article or through supplementary data files.
The regentrans R package can be downloaded from GitHub: https://github.com/Snitkin-Lab-Umich/regentrans/
The manuscript figures are generated from regentrans example data and can also be found on GitHub: https://github.com/Snitkin-Lab-Umich/regentrans/tree/master/vignettes/manuscript_figures
The example data used in the package and manuscript is from BioProject accession no. PRJNA415194. The metadata corresponding to these sequences can be found on the SRA Run Selector (isolate column) and as example data in the regentrans package.

https://github.com/Snitkin-Lab-Umich/regentrans/

## Authors and contributors

All authors developed methodology and reviewed and edited the manuscript. SH, ZL, and ESS authors conceptualized the project. SH and ZL curated the data, designed the software, and visualized the results. ZL, JW, and ESS acquired funding. ZL and ESS wrote the original manuscript draft. ESS supervised the project.

## Conflicts of interest

The authors declare that there are no conflicts of interest.

## Funding information

ESS received support from the National Institutes of Health under Grant No. 1R01AI148259-01. ZL received support from the National Science Foundation Graduate Research Fellowship Program under Grant No. DGE 1256260. JW received support from the Canadian Institutes of Health Research fellowship Grant No. 201711MFE-396343-165736 and the Michigan Institute for Clinical and Health Research (MICHR) Postdoctoral Translational Scholars Program (PTSP). Any opinions, findings, and conclusions or recommendations expressed in this material are those of the authors and do not necessarily reflect the views of the National Science Foundation. The funding bodies had no role in the design of the study or collection, analysis, and interpretation of data, or in writing the manuscript.

## Ethical approval

N/A

## Consent for publication

N/A

## Acknowledgements

We would like to thank Emily Benedict for testing out the alpha version of our package.

## Notes

### Competing Interest Statement

The authors have declared no competing interest.

### Author Declarations

The original study was reviewed and approved by the Institutional Review Board of the University of Pennsylvania with a waiver of informed consent.

### Summary of Updates

Updated text and figures

